# On the treatment effect heterogeneity of antidepressants in major depression. A Bayesian meta-analysis

**DOI:** 10.1101/2020.02.20.19015677

**Authors:** Constantin Volkmann, Alexander Volkmann, Christian A. Müller

## Abstract

**Background:** The average treatment effect of antidepressants in major depression was found to be about 2 points on the 17-item Hamilton Depression Rating Scale, which lies below clinical relevance. Here, we searched for evidence of a relevant treatment effect heterogeneity that could justify the usage of antidepressants despite their low average treatment effect.

**Methods:** Bayesian meta-analysis of 169 randomized, controlled trials including 58,687 patients. We considered the effect sizes log variability ratio (lnVR) and log coefficient of variation ratio (lnCVR) to analyze the difference in variability of active and placebo response. We used Bayesian random-effects meta-analyses (REMA) for lnVR and lnCVR and fitted a random-effects meta-regression (REMR) model to estimate the treatment effect variability between antidepressants and placebo.

**Results:** The variability ratio was found to be very close to 1 in the best fitting models (REMR: 95% HPD [0.98, 1.02], REMA: 95% HPD [1.00, 1.02]). The between-study variance τ^2^ under the REMA was found to be low (95% HPD [0.00, 0.00]). Simulations showed that a large treatment effect heterogeneity is only compatible with the data if a strong correlation between placebo response and individual treatment effect is assumed.

**Conclusions:** The published data from RCTs on antidepressants for the treatment of major depression is compatible with a near-constant treatment effect. Although it is impossible to rule out a substantial treatment effect heterogeneity, its existence seems rather unlikely. Since the average treatment effect of antidepressants falls short of clinical relevance, the current prescribing practice should be re-evaluated.

## Introduction

Depression is one of the most frequent psychiatric disorders and poses a major burden for individuals and society; it affects more than 300 million people worldwide and is ranked as the single largest contributor to disability [1]. The first-line treatment usually consists of psychotherapy and/or pharmacotherapy with antidepressant drugs [2, 3]. Within the last decades, the number of prescriptions of antidepressants has continuously increased in several regions of the world [4, 5]. However, whether antidepressants are effective in the treatment of major depression has been a highly controversial debate for many years [6-9]. A recent meta-analysis by Cipriani et al. comprising 522 randomized, controlled trials (RCTs) of 21 antidepressants in 116□477 participants reported that all antidepressants were more effective than placebo in reducing depressive symptoms [10]. In contrast, the authors of a recent re-analysis criticised this meta-analysis for not taking into account several biases, such as publication bias [12]. They concluded that “the evidence does not support definitive conclusions regarding the efficacy of antidepressants for depression in adults, including whether they are more efficacious than placebo for depression”.

Albeit these contradictory conclusions, both analyses used the same dataset. The so-called average treatment effect, which measures the difference in mean outcomes between active and control group, was about 2 points on the 17-item Hamilton Depression Rating Scale (HAMD-17) [13] in this dataset [12]. According to Leucht et al. [14], a reduction of up to 3 points on the HAMD corresponds to “no change” in the Clinical Global Impressions - Improvement Scale (CGI-I) [15] and the assumed threshold of clinical significance is 7 points [16]. Thus, a reduction of 2 points on the HAMD is not detectable by the treating physician and is presumably clinically irrelevant.

Crucially, Munkholm et al. [12] reported the average treatment effect as an outcome parameter, whereas Cipriani et al. [10] reported the odds ratio (OR) of “response rates”, signifying the fraction of patients crossing the rather arbitrary threshold of 50% in symptom reduction (“responders”). This approach translates into a number-needed-to-treat (NNT) of around 8-10 for “response” [17].

### Categorisation of continuous variables

Categorising patients into “responders” and “non-responders” based on crossing an arbitrary threshold on a continuous scale has frequently been criticised by statisticians as it may lead to an artificial inflation of the measured treatment effect and to a loss of power [18, 19]. It may create the illusion of a subgroup of patients that benefit particularly well from a given treatment where none exists. However, a NNT of 8 is compatible with every patient having the exact same treatment effect of 2 points on the HAMD-17 [12, 20]. Only if a substantial so-called treatment effect heterogeneity exists, meaning that there are true “responders” and “non-responders”, the calculation of response rates may be legitimate and the average treatment effect may not be an appropriate outcome measure. However, in the absence of clear evidence for a relevant treatment effect heterogeneity, the average treatment effect is the best predictor of the individual treatment effect [20, 21].

### Treatment effect heterogeneity

Treatment effect heterogeneity describes the extent to which a treatment might affect different individuals differentially. In other words, some patients may benefit a lot, others may be harmed by a given treatment, possibly resulting in a null finding when only considering the average treatment effect in clinical trials. However, the existence of a clinically relevant treatment effect heterogeneity, albeit widely believed and intuitively plausible, has not been shown yet.

### Investigation of treatment effect heterogeneity

Simply labeling patients as “responders” and “non-responders” based on crossing an arbitrary threshold on a continuous outcome scale is not a valid way to investigate variation in individual treatment effect [22]. In order to assess treatment effect heterogeneity from the data of parallel group trials, the comparison of variances between the active and the control condition has been proposed [22, 23]. Here, an increase in variance in the active group might be a signal of a variation in the individual treatment effect [22].

Following a recent publication by Winkelbeiner et al. [21], analyzing differences in variances in 52 randomized, placebo-controlled antipsychotic drug trials, the present analysis aimed to assess the evidence for individual antidepressant drug response using the open dataset of the largest meta-analysis of the efficacy of antidepressants in major depressive disorder [10].

Here, we addressed the following research question: What is the evidence for a relevant treatment effect heterogeneity of antidepressants in the treatment of depression that justifies their usage despite the lack of a clinically relevant average treatment effect?

## Methods

### Data Acquisition

We obtained the dataset of the meta-analysis by Cipriani et al. [10] from the Mendeley database (https://data.mendeley.com/datasets/83rthbp8ys/2). This study included all RCTs comparing 21 antidepressants with placebo or another active antidepressant as oral monotherapy for the acute treatment of adults (≥18 years old and of both sexes) with a primary diagnosis of major depressive disorder according to standard operationalized diagnostic criteria (Feighner criteria, Research Diagnostic Criteria, DSM-III, DSM-III-R, DSM-IV, DSM-5, and ICD-10). For further details on the inclusion criteria and study characteristics, see the original study [10].

### Data extraction and processing

Of the total of 522 studies we kept the 304 that included a placebo arm. We excluded all studies for which the reported endpoint did not represent the change from baseline, leaving us with a total of 169 studies for the analysis (see PRISMA flow diagram, supplementary data, figure 1). We extracted both the mean and the standard deviation of pre- and post-treatment outcome difference scores (the “response”). The studies included in the data set comprised 8 different depression scales, namely HAMD-17, HAMD-21, HAMD-24, HAMD unspecified, HAMD-29, HAMD-31, Montgomery–Åsberg Depression Rating Scale (MADRS) and IDS-IVR-30 [25, 26]. Studies with different treatment arms were aggregated according to the recommendation of the Cochrane Collaboration [27]. In this manuscript, we define response as pre-post-difference of a given outcome scale.

**Figure 1:**
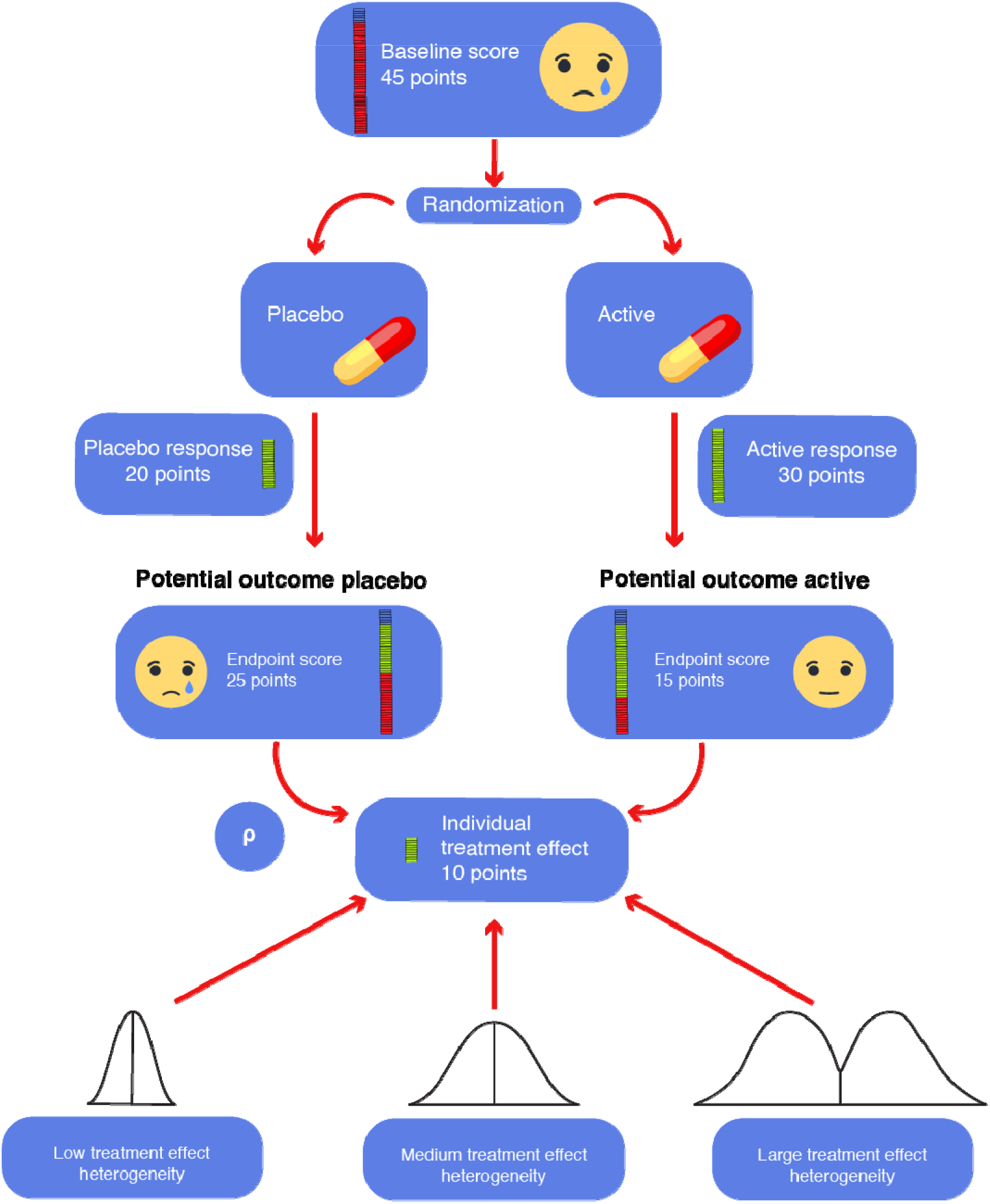
Visualization of a hypothetical patient in a randomized placebo-controlled trial. The patient is randomized to either the placebo or the active arm, corresponding to two hypothetical “potential outcomes”. Only one of which can ever be observed, as a single patient cannot receive both placebo and the active intervention at the same time. The difference between the two potential outcomes corresponds to the “individual treatment effect” of the intervention (here, a clinically relevant difference of 10 HAMD-17 points). The individual treatment effect is unobservable and can be imaged to be drawn from hypothetical distributions of the treatment effect. The variance of this distribution corresponds to the treatment effect heterogeneity. The factor ρ is the correlation between the placebo response and the individual treatment effect. Here, we assume that a given patient has a fixed individual treatment effect.

### Statistical Analysis

We considered two different effect size statistics as suggested by Nakagawa et al. [28] to analyze the difference in variability of active and placebo response.

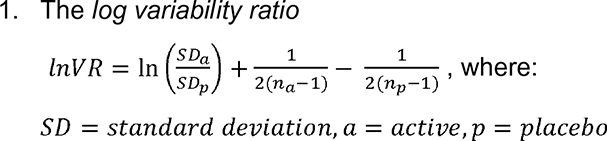

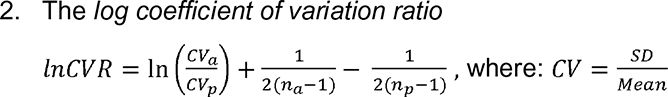

These two effect sizes differ in the way they account for differences in means between the active and the placebo group. Whereas lnVR assumes no correlation between concurrent changes in mean response and standard deviation of response, lnCVR measures differences in variability between groups after accounting for differences in mean response. If the active and placebo arms have equal variance, a VR (or CVR) of 1 would be expected. A value greater than 1 indicates a larger variability in the active group.

A variability ratio that substantially differs from 1 implies a considerable treatment effect heterogeneity. Conversely, a VR of around 1 is compatible with a near-constant treatment effect but does not exclude the existence of treatment effect heterogeneity. It should be noted, that it is impossible to disprove the existence of a subgroup with a substantially greater than average effect. However, the magnitude of the treatment effect heterogeneity can be bounded by the distance of the variability ratio VR from the value 1.

All statistical analyses were carried out in the programming language Python (version 3.7) and the probabilistic programming language Stan (with pystan version 2.18.1.0 as a Python interface). We used a Bayesian approach to fit all our models using weakly informative priors. Firstly, we used a Bayesian random-effects meta-analyses (REMA) for the two effect statistics lnVR and lnCVR. Secondly, we used a Bayesian random-effects meta-regression (REMR) to fit the lnVR effect statistic with the natural logarithm of the response ratio (lnRR) as a regressor [28], which is defined as:

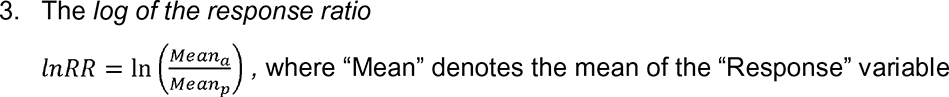

An additional complexity in our analysis, as compared to recent analyses [21, 28, 29], came from the fact that our data set contained several different depression scales (several versions of the HAMD and the MADRS, see supplementary figure 2). For our analysis we made the assumption that these different scales are (locally) linearly transformable into each other. This assumption is well supported by the literature [30]. Fortunately, the lnVR and lnCVR effect statistics are invariant under linear transformations of the outcome scale.

### Random-effects meta-analysis (REMA)

We applied a Bayesian random-effects meta-analysis in order to estimate the effect sizes ES=lnVR, lnCVR. For the REMA, the following model was applied, where µ equals the “true” mean of the effect size. Finally, η represents the between-study-variance.

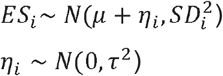

We specified the following weakly-informative hyper-priors:

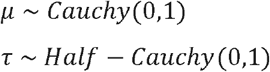

### Random-effects meta-regression (REMR)

This approach is a “contrast-based” version of the “arm-based” meta-analysis in Nakagawa et al. [28] which models the log of the standard deviation of the outcome directly in a multi-level meta-regression. For the REMR, the following model was applied, where µ equals the “true” mean of lnVR over all studies and X the “true” value of lnRR, if we account for measurement error. The variable β is the regression coefficient for X and thus signifies the degree of linear association between lnVR and lnRR. Finally, η represents the between-study-variance.

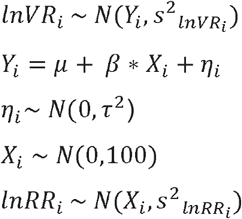

We specified the following weakly-informative hyper-priors:

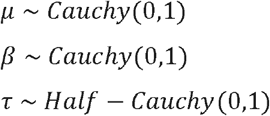

### Simulation experiments

For each simulation, the response under placebo and the response under treatment were simulated for 1000 patients. The response under placebo was drawn from a right skewed distribution with mean and standard deviation of 8.8 and 7.7 points on the HAMD-17 scale (based on Cipriani data [10]), respectively. For each patient, an outcome under treatment was computed from a mixed Gaussian distribution with a given SD_TE,_ where the outcome under placebo and the individual treatment effect were required to be correlated by the correlation coefficient ρ.

This yielded a potential outcome under placebo and a potential outcome under active treatment for every patient with a corresponding individual treatment effect (see figure 1 for illustration). Half of the patients were then randomly selected for treatment, the other half was assigned to placebo. Note that only one of these two outcomes can be observed in a real experiment.

## Results

### Study selection

As mentioned above, we included 169 placebo-controlled studies that reported mean and standard deviation of change in depression scores. These studies included data on 58,687 patients treated with 21 different antidepressants.

### Correlation between mean and standard deviation of depression scores

In order to identify the more appropriate effect size (VR or CVR), we investigated the linear association between the logarithm of the mean response scores and the logarithm of their standard deviation using a varying intercept model, where the intercepts were allowed to vary between studies with different depression scales. Fitting a Bayesian varying intercept regression model with measurement error with lnMean as independent variable and lnSD as dependent variable, we get a posterior mean for the slope coefficient of 0.10 with a 95% HPD (highest probability density) interval of [0.04, 0.16]. This can be interpreted as a weak correlation between lnMean and lnSD. We remark that simply computing the correlation of the two quantities without paying attention to the correct weighting and the different scales in the data would yield an overestimated slope coefficient of 0.25 (see supplementary table 1).

### Log variability ratio (lnVR) and log coefficient of variation (lnCVR) models

In order to estimate the difference in variability between antidepressant and placebo response, we modelled the lnVR effect size using a Bayesian random effects model as heterogeneity between studies may be expected. The posterior mean estimate for the variability ratio was 1.01, with the 95 % highest posterior density (HPD) interval ranging from 1.00 to 1.02. The lnVR effect size assumes no correlation between lnMean and lnSD and may give biased results if such a correlation exists. In the presence of a positive correlation between mean and standard deviation, Nakagawa et al. [28] suggest that the lnCVR may be the more appropriate effect size to investigate the difference in variability between the active and control. The lnCVR REMA showed a reduction in the coefficient of variation in the active versus the placebo group (posterior mean estimate for CVR: 0.82, 95% HPD [0.80,0.84]).

### Random-effects meta-regression

Finally, we used a Bayesian random effects meta-regression (REMR). The advantage of this model over the lnVR and lnCVR meta-analyses is that we are not forced to make rigid assumptions about the association between the lnMean and lnSD, as the strength of this relationship is estimated directly from the data. Fitting this model, we obtained posterior statistics for the μ and β coefficients. The posterior mean estimate for e^μ^ was 1.00 (95% HPD [0.98,1.02]) and that for β 0.04 (95% HPD [-0.03,0.12]), where we can (roughly, up to measurement error and random noise) interpret the coefficients as follows:

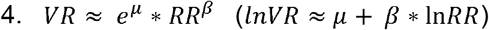

Note that the lnVR REMA corresponds to a lnVR REMR with a β coefficient set to 0, whereas the lnCVR REMA corresponds to a lnVR REMR with a β coefficient set to 1. The REMR model learns the β coefficient and its posterior HDP interval is equal to 0.04 [-0.03, 0.12] suggesting that the lnVR REMA is a more appropriate model than the lnCVR REMA.

### Between-study heterogeneity

The between-study variance τ^2^ under the REMA was found to be low for both lnVR (95% HPD [0.00,0.00]) and lnCVR (95% HPD for τ^2^ [0.00,0.01]). Indeed, applying a fixed effects model instead of the REMA for the purpose of sensitivity analysis yielded similar results for the overall mean estimates of lnVR and lnCVR.

### Performance comparison of the different models

In order to compare the performance of the different models applied, we used the so-called *widely applicable information criterion* (WAIC). This method estimates the pointwise prediction accuracy of fitted Bayesian models. Here, higher values of WAIC indicate a better out-of-sample predictive fit (“better” model). We refer to Vehtari et al. [31] for more details on WAIC. Figure 4 shows the logWAIC for the different models.

**Figure 2:**
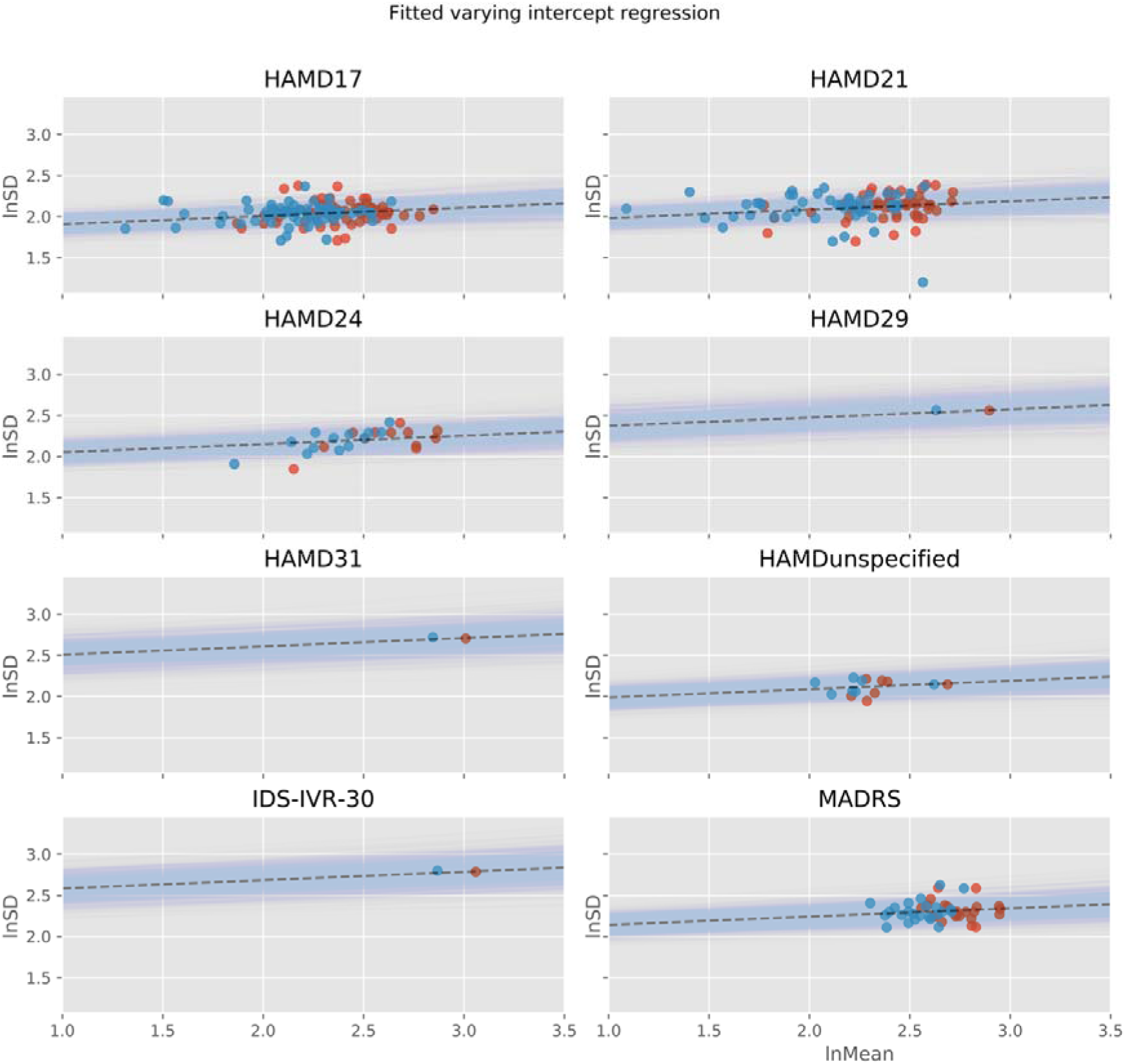
Linear association between lnMean and lnSD using a varying intercept model, where the intercepts are allowed to vary between studies with different depression scales. Red dots represent active groups, blue dots represent placebo groups.

**Figure 3:**
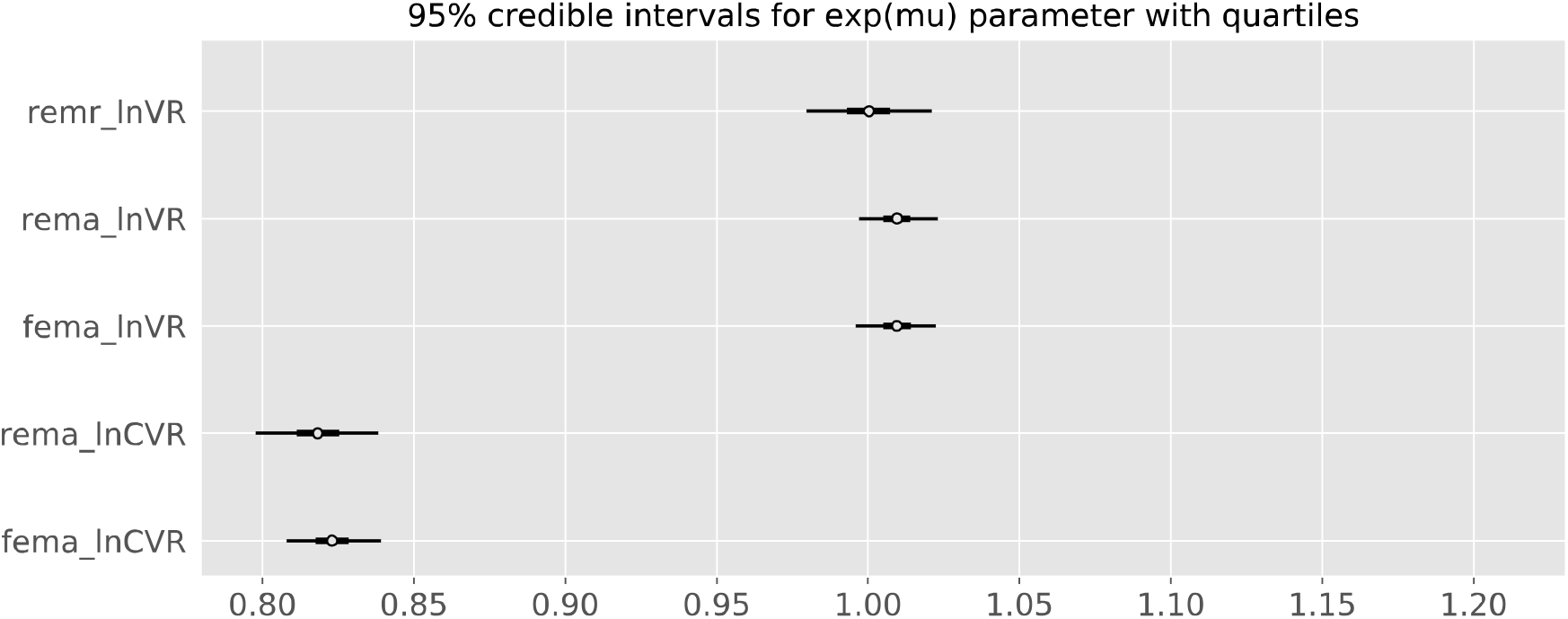
Posterior credible intervals for the exp(µ) parameter for the different models. REMA: random-effects meta-analysis. FEMA: fixed-effects meta-analysis. REMR: random-effects meta-regression. Note that the results are very similar for the REMR and the lnVR meta-analyses.

**Figure 4:**
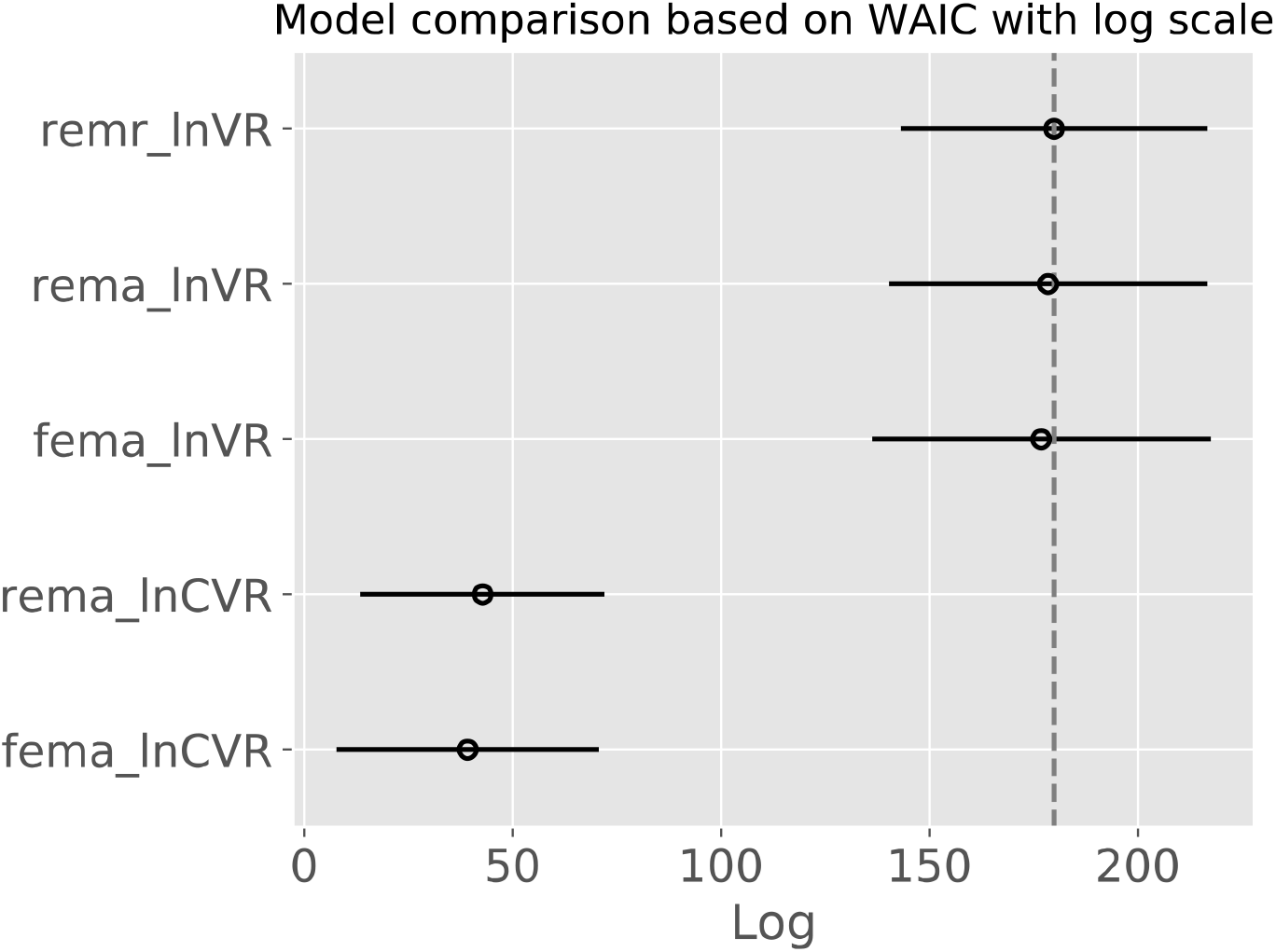
*Widely applicable information criterion* (WAIC) depicted on a logarithmic scale. Higher values signify a better predictive fit of the underlying model. Bars indicate standard errors. REMA: random-effects meta-analysis. FEMA: fixed-effects meta-analysis. REMR: random-effects meta-regression.

We observed that the lnVR REMA and the lnVR REMR outperformed the lnCVR REMA with respect to the WAIC. The difference between the lnVR REMA and the lnVR REMR showed comparable performance with respect to the WAIC.

### Upper bound on the treatment effect heterogeneity

In order to investigate the compatibility of different assumptions regarding the treatment effect with the measured variability ratio, we use the following equation that was derived in [32]:

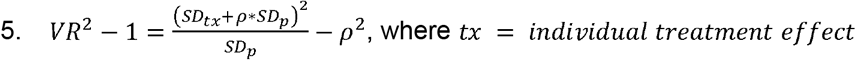

Note that the variable ρ signifies the degree of correlation between the treatment effect tx and the response under placebo p (see figure 1) and is unobservable. Assuming that VR^2^ −1 is smaller than some number ε, the above equation implies that:

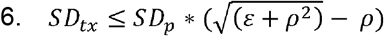

Using the Cipriani et al. dataset [10] we can estimate SD_p_ to be equal to around 7.66 on the HAMD-17 scale. From our meta-analysis of the lnVR effect statistic, we have that the 95th percentile of the posterior distribution of e^μ^ is 1.02. This implies the following inequality:

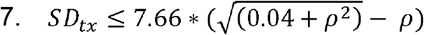

This inequality tells us that if we assume any value ρ ∈ [−1, 1] (the correlation between the treatment effect and the response under placebo), we get an upper bound on the standard deviation of the treatment effect as above.

### Which distributions of the treatment effect are possible for a VR of nearly 1

Based on the above-mentioned formula 7, table 1 depicts different magnitudes of treatment effect heterogeneity compatible with a VR of 1.02, which is the 95th percentile of our VR estimate.

**Table 1:** Assuming a VR of 1.02, a SD_p_ of 7.66 (based on Cipriani [10]) and different correlation coefficients ρ between the response under placebo and the treatment effect. Left column (“any distribution”): Upper bounds for the standard deviation of the treatment effect. Right column (“dichotomous response”): Patients are either “non-responders” with a treatment effect of 0, or “responders” with the responder treatment effect. For a given correlation coefficient ρ, there is one possible solution for this. TE: treatment effect.

|  | any distribution |  | dichotomous response |  |
| --- | --- | --- | --- | --- |
|  | SD(TE) | % > 7 HAMD | responder TE | % responder |
| $\rho$ | | | | |
| -1.0 | 15.5 | 37.3 | 20.5 | 9.7 |
| -0.8 | 12.4 | 34.4 | 24.3 | 8.2 |
| -0.6 | 9.4 | 29.8 | 29.8 | 6.7 |
| -0.4 | 6.5 | 22.1 | 38.1 | 5.2 |
| -0.2 | 3.7 | 8.9 | 51.9 | 3.9 |
| 0.0 | 1.5 | 0.1 | 72.2 | 2.8 |
| 0.2 | 0.6 | 0.0 | 86.2 | 2.3 |
| 0.4 | 0.4 | 0.0 | 91.6 | 2.2 |
| 0.6 | 0.3 | 0.0 | 94.1 | 2.1 |
| 0.8 | 0.2 | 0.0 | 95.5 | 2.1 |
| 1.0 | 0.2 | 0.0 | 96.3 | 2.1 |

The left column (“any distribution”) of the table depicts the upper bound for the standard deviation of the treatment effect (the treatment effect heterogeneity) with a VR of 1.02. The upper bound for the treatment effect heterogeneity depends upon the population-level correlation ρ between the “response under placebo” and the “individual treatment effect” (see figure 1). The results can be interpreted as follows: assuming a correlation ρ between the individual treatment effect and the response under placebo of a given value in the table, we are 95% sure (under the assumptions of the meta-analytic model (REMA) of lnVR) that the standard deviation of the treatment effect variable is smaller than the corresponding value of the second column in the table. Furthermore, assuming a minimally clinically relevant effect of 7 points on the HAMD-17 scale, the third column tells us the percentage of patients with a medication effect at least as large for the largest possible standard deviation within the 95% credible interval. Note that these results are independent of the distribution of the treatment effect (normal, binormal, etc.), as these results were derived analytically from the above-mentioned formula (see formula 7).

The right column assumes a dichotomous treatment effect (“responder”, “non-responder”). Here, “non-responders” are assumed to have treatment effect of 0 (placebo response = antidepressant response), whereas “responders” have a fixed treatment effect > 0. For a given VR of 1.02, the percentage of “responders” and their respective “responder treatment effect” depend upon the intra-individual correlation ρ between the potential outcome placebo response and individual treatment effect.

These results show that, contrary to intuition, a variability ratio of 1.02 is (theoretically) compatible with a standard deviation of the treatment effect between 0 and 15.5. Conversely, a reduction in the variability in the treatment group is compatible with a substantial treatment effect heterogeneity if the response under placebo is correlated with the individual treatment effect (see simulations in supplementary file, figures 8 and 9).

### Simulations

We conducted simulation experiments in order to illustrate the compatibility of a VR of 1.02 with different degrees of treatment effect heterogeneity. For a large treatment effect heterogeneity, the individual treatment effect was drawn from a distribution with an (arbitrarily chosen) standard deviation SD_TE_ = 6.5 HAMD-17. For this SD_TE_, the response under placebo and the individual treatment effect have to be correlated by the correlation factor ρ = - 0.4, in order for the VR to credibly remain at or below 1.02 (see table 1). Figure 5 depicts the change scores of 1000 patients under placebo (blue) and under active treatment (red). Here, positive values denote an improvement of the depression severity.

**Figure 5:**
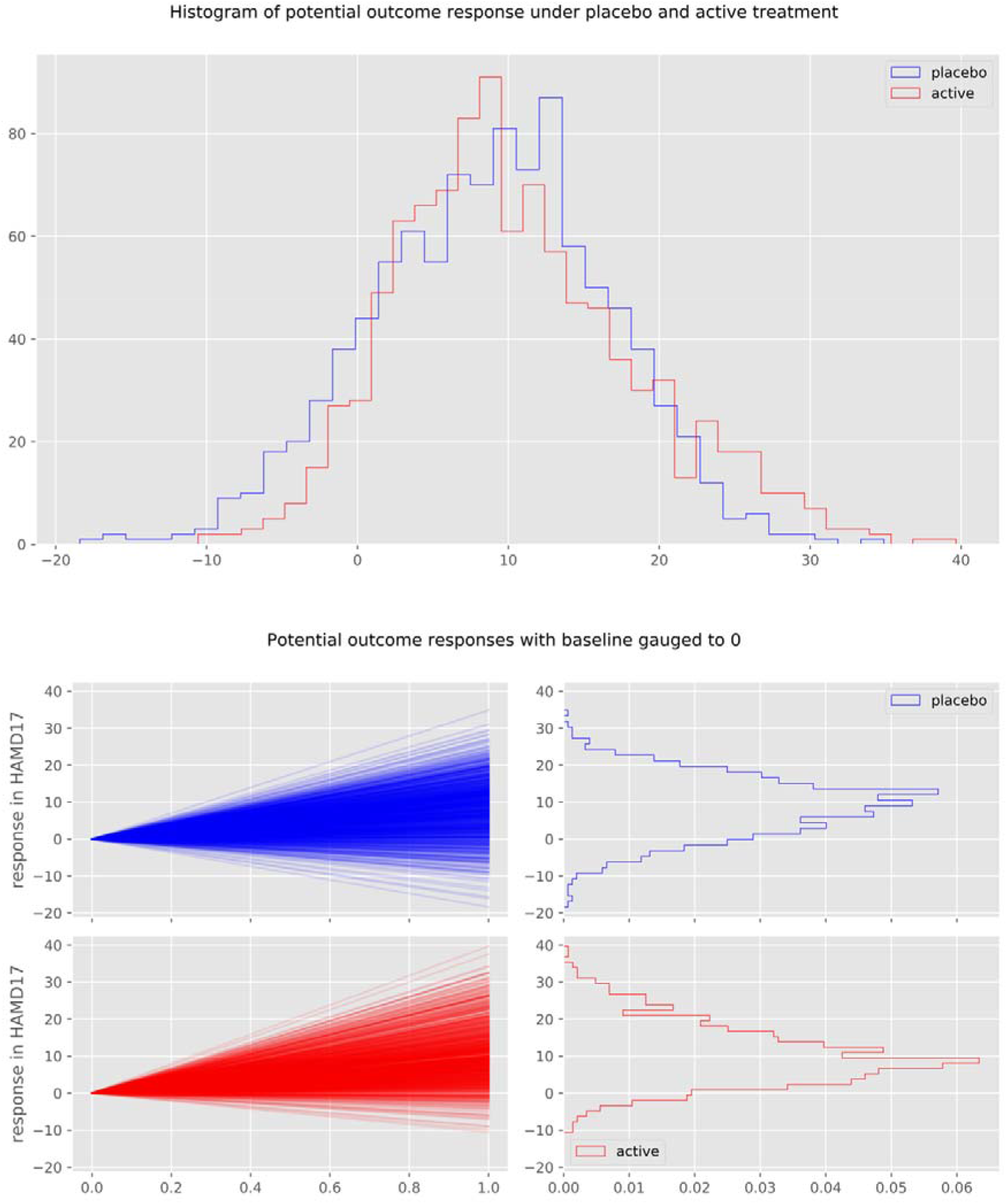
Change score of 1000 simulated patients under placebo (blue) and under active treatment (red) for ρ = - 0.4, SD_TE_ = 6.35 HAMD-17 points and VR = 1.02. Note, that in this particular simulation, the SD_TE_ is not exactly equal to 6.5, as all simulations contain random processes.

Conversely, if the correlation factor ρ is equal to 0, a large treatment effect heterogeneity with SD_TE_ = 6.5 would yield a VR in the magnitude of 1.3. For the VR to be credibly lower than 1.02 and the response under placebo and the individual treatment effect to be uncorrelated (ρ = 0), the treatment effect heterogeneity has to be low. Supplementary figures 6 and 7 depict the results of such a simulated experiment. Here, the treatment effect heterogeneity was imputed to be SD_TE_ = 1.5 points on the HAMD-17 (derived from table 1). A VR closer to 1 would yield an even smaller treatment effect heterogeneity.

Figure 6 shows the magnitude of the individual treatment effect of 100 individuals of this simulation experiment. The values on the left (x = 0.0) denote the response under placebo of all 100 patients, the values on the right (x = 1.0) represent the response under active treatment and the difference between the two values corresponds to the individual treatment effect of a patient. As can be seen in the figure, for a large treatment effect heterogeneity to be compatible with a VR of 1.02, the response under placebo and the individual treatment effect have to be correlated. Specifically, patients that would remain unchanged under placebo (response close to 0 at x = 0.0) have a larger benefit from active medication (higher density of blue slopes) than patients that would have improved under placebo (response above 0 at x = 0.0). For some patients to benefit substantially, however, other patients have to be harmed by the medication (red slopes).

**Figure 6:**
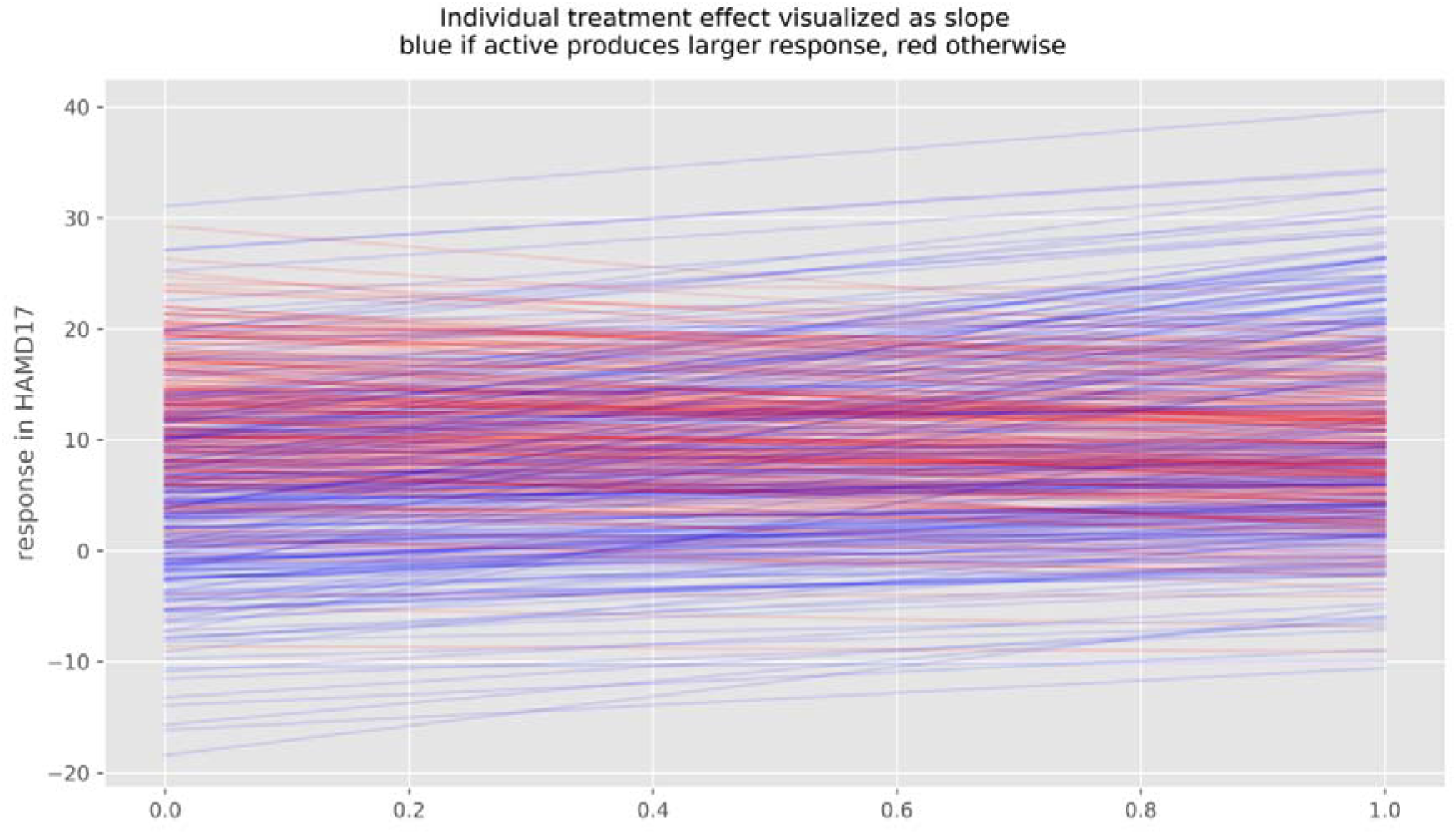
Potential outcomes and individual treatment effect of 100 simulated patients. Value at x = 0 depicts response under placebo, value at x = 1 response under active treatment. Slopes represent individual treatment effect, which varies substantially in this simulation. Blue lines indicated improvement under active treatment, red lines deterioration.

## Discussion

The efficacy of antidepressants in the treatment of major depressive disorder has been the topic of an ongoing debate for years in the psychiatric community and the public [6-9]. In a recent re-analysis [12] of a network meta-analysis [10], the average treatment effect of antidepressants was found to be about 2 points on the HAMD-17 scale, which is almost undetectable by clinicians [14] and clearly lies below the assumed minimally clinically relevant effect of 7 points [16]. In addition, it should be noted that relevant biases may have led to an overestimation of the drugs’ efficacy [12, 33]. Jakobsen et al. recently concluded that, based on current evidence, antidepressants should not be used in adult patients with major depressive disorder [34].

### Evidence for treatment effect heterogeneity

On the basis of these facts, the question arises as to why these compounds are considered to be effective, nevertheless. The main reason for this might be the assumption of a substantial treatment effect heterogeneity, meaning that subpopulations of patients exist that benefit substantially more than average from the medication. If the treatment effect heterogeneity is low, no patient would have a clinically relevant benefit. In the face of the well documented harms and side effects, antidepressants may then not be considered useful in the treatment of major depression.

Albeit widely believed and putatively observed in clinical routine, substantial differences in the individual treatment effect of antidepressants have not been shown to exist yet. A “responder”, usually defined as someone crossing an arbitrary threshold of symptom severity, is a person who was observed to improve and not necessarily caused by the medication to get better. Even constant treatment effects lead to differences in observed response rates, creating the illusion of a differential treatment effect, where none exists [20]. Therefore, the frequently performed calculation of response rates is misleading and seems inappropriate to answer the question of treatment effect heterogeneity.

This work aimed to estimate the treatment effect heterogeneity of antidepressants in the treatment of major depressive disorder using a large dataset of a recent network meta-analysis [10]. To this end, we applied the effect size statistics lnVR and lnCVR suggested by Nakagawa et al. [28], using a Bayesian random-effects meta-analytical approach (REMA) and fitted a multi-level meta-regression (REMR) model to estimate the treatment effect variability between antidepressants and placebo. Both the lnVR REMR and the lnVR REMA, which were found to outperform the lnCVR REMA, showed that the variability ratio was very close to 1 (REMR: 95% HPD [0.98, 1.02], REMA: 95% HPD [1.00, 1.02]), perfectly compatible with a near-constant effect of antidepressants on depression severity. These findings are in line with those of a recently published meta-analysis of antidepressants using the same dataset [35].

### Methodological aspects

In order to determine the variability of the treatment response, the correlation between the mean and standard deviation of the underlying measuring scale has to be taken into account. The lnVR and the lnCVR effect sizes naively assume a slope coefficient of 0 and 1, respectively. In other words, how much of the (logarithmic) difference in variances is explained by the difference in (logarithmic) means. Both scales may thus give biased results, if the true slope coefficient differs from the one assumed.

By applying a varying intercept model, taking into account the occurrence of different depression scales, we could show that the correlation between (logarithmic) mean and (logarithmic) standard deviation is of a small magnitude (slope coefficient = 0.10), indicating that the lnVR is a more appropriate measure as opposed to the lnCVR. A regression over all depression scales yields a slope coefficient of 0.25, which is 2.5 x as large as our estimate.

When simply conducting a significance test for the existence of such correlation, the lnCVR effect size would appear to be the appropriate measure, leading to the incorrect conclusion of a substantially reduced variability in the active arm. It is important to note that a VR (or CVR) sufficiently smaller than 1 is in fact evidence of relevant treatment effect heterogeneity (see supplementary figure 8 and 9). Therefore, considering the lnCVR as the main outcome would lead to the opposite conclusion of substantial treatment effect heterogeneity [36].

Our work adds accuracy to the existing literature, as we developed a generalized model (REMR) that incorporates the slope coefficient for the correlation between mean and standard deviation directly from the data. This approach yielded a mean estimate for the VR of 1.00 (95% HPD [0.98,1.02]), again compatible with a near-constant effect.

We applied the WAIC in order to estimate the predictive power of our models. This effect measure showed that the models using the lnVR effect size had a better out of sample predictive power than the models using the lnCVR effect size.

### Upper bound for the treatment effect heterogeneity

As a relevant treatment effect heterogeneity cannot be ruled out even with a variability ratio near 1, we aimed at estimating the upper bound in treatment effect variation compatible with our results. We were able to analytically derive an inequality that provides an upper bound for the treatment effect heterogeneity, taking into account a possible correlation between the placebo response and the individual treatment effect. We could show that a VR of 1.02 (the upper bound of the 95% HPD interval of the REMR) is theoretically compatible with a standard deviation of the treatment effect between 0 and 15.5 points on the HAMD-17 scale, translating into a maximum of 37% of patients with an individual treatment effect of more than 7 points on the HAMD-17 scale.

However, such a large standard deviation and hence treatment effect heterogeneity would require the treatment effect and the response under placebo of a patient to be strongly and negatively correlated in order to be compatible with a VR of 1.02. If no such correlation exists, the treatment effect heterogeneity would be negligibly small (SD_TE_ = 1.5 HAMD-17 points, 0.1% of patients benefitting more than 7 points on the HAMD-17 scale).

As only one outcome per patient (either under placebo or under active treatment; figure 1) can be measured in a real experiment, the true correlation ρ between the response under placebo and the treatment effect cannot be derived from RCT data.

### How should these results be interpreted

The VR is a measure that can potentially detect evidence for subgroups that benefit (substantially) more than average from an intervention. A VR that differs substantially from 1 is evidence of such subgroups (of large treatment effect heterogeneity), while a VR near 1 is compatible with both a small and a large treatment effect heterogeneity. A VR of exactly 1 (which is the mean-estimate of our REMR model) would be proof of a constant treatment effect. It is, however, impossible to ever prove identity, as we can never reach an uncertainty of 0 (credible interval with width of 0). Furthermore, an exactly constant treatment effect seems impossible also from a theoretical point of view. So how should a VR of 1 (95% HPD [0.98,1.02]) be interpreted? For this, consider the following illustration:

*Hypothesis 1 (H1):* The treatment effect heterogeneity is close to 0 (e.g. 99% of patients have an individual treatment effect of 1 to 3 HAMD points).

*Hypothesis 2 (H2):* The treatment effect heterogeneity is greater than in H1.

There are now three possibilities:

1. H1 is true and VR ≈ 1 (very close to 1, e.g. 0.98 to 1.02)
2. H2 is true and VR ≈ 1
3. H2 is true and VR ≠ 1 (not very close to 1)

Our results indicate that VR ≈ 1. We can thus rule out one of the three possibilities, namely a large treatment heterogeneity combined with a VR ≠ 1. From a Bayesian perspective, the probability of H1 being true increases, while that of H2 being true decreases. How we now regard the probability of H1 or H2 being true depends on how plausible we considered these scenarios to begin with (the prior probabilities).

In order for H2 to be true and the VR being close to 1, strong assumptions regarding the correlation between the placebo response and the individual treatment effect of antidepressants are necessary. Specifically, those patients whose depression severity would remain unchanged under placebo would need to have the strongest antidepressant medication effect. If this were the case, we might expect patients with certain features (such as chronic depression) to benefit substantially more than average from antidepressants. Since no such subpopulations have been identified to date, such a correlation seems unlikely. If no such correlation is assumed, a VR of 1.02 indicates a low degree of treatment effect heterogeneity.

## Conclusion

By applying a multiple level Bayesian regression model and simulations, this work could show that the published data on antidepressants in the treatment of major depression is compatible with a near-constant treatment effect, which is also the simplest explanation for the observed data. Although is not possible to rule out a substantial treatment effect heterogeneity using summary data from RCTs, we could show that a substantial treatment effect heterogeneity is only compatible with the published data under strong assumptions that seem rather unlikely. Until the existence of benefiting subgroups has been demonstrated prospectively, the average treatment effect is the best estimator for the individual treatment effect. Since the average treatment effect of antidepressants probably falls short of clinical relevance, the current prescribing practice in the treatment of major depression should be critically re-evaluated.

## Data Availability

All data and the python code are available online.

https://data.mendeley.com/datasets/83rthbp8ys/2

https://github.com/volkale/advr

## Python code

https://github.com/volkale/advr

## Statement of Ethics

The authors have no ethical conflicts to disclose.

## Disclosure Statement

CAM received consulting fees from Silence Therapeutics, outside the submitted work. The other authors declared no competing interest. All authors declare no other relationships or activities that could appear to have influenced the submitted work. No funder had any role in: the design and conduct of the study; collection, management, analysis, and interpretation of the data; preparation, review, or approval of the manuscript; and decision to submit the manuscript for publication.

## Funding Sources

The authors received no specific funding for this work.

## Author Contributions

Study idea and design: CV and CAM. Data extraction, statistical analyses, visualizations and simulation experiments: AV and CV. Mathematical modelling, code implementation and derivation of formula: AV. All authors contributed to drafting the manuscript. All authors provided a critical review and approved the final paper.

## Notes

### Funding Statement

No funding.

## References

1. WHO. Depression and Other Common Mental Disorders: Global Health Estimates. 2017; Available from: https://apps.who.int/iris/bitstream/handle/10665/254610/WHO-MSD-MER-2017.2-eng.pdf.

2. DGPPN, S3-Leitlinie und Nationale VersorgungsLeitlinie (NVL) Unipolare Depression, 2. Auflage. 2015.

3. NICE, Depression in adults: recognition and management. 2009. [28.08.2019] Available from: https://www.nice.org.uk/guidance/cg90/chapter/1-Guidance#treatment-choice-based-on-depression-subtypes-and-personal-characteristics

4. BMJ, NHS prescribed record number of antidepressants last year, in BMJ. 2019. 1508.

5. Kantor, E.D., et al., Trends in Prescription Drug Use Among Adults in the United States From 1999-2012. JAMA, 2015. 314(17): p. 1818–31.

6. Moncrieff, J. and I. Kirsch, Efficacy of antidepressants in adults. BMJ, 2005. 331(7509): p. 155–7.

7. Fountoulakis, K.N. and H.J. Moller, Efficacy of antidepressants: a re-analysis and re-interpretation of the Kirsch data. Int J Neuropsychopharmacol, 2011. 14(3): p. 405–12.

8. Davis, J.M., et al., Should we treat depression with drugs or psychological interventions? A reply to Ioannidis. Philos Ethics Humanit Med, 2011. 6: p. 8.

9. Gotzsche, P.C., Why I think antidepressants cause more harm than good. Lancet Psychiatry, 2014. 1(2): p. 104–6.

10. Cipriani, A., et al., Comparative efficacy and acceptability of 21 antidepressant drugs for the acute treatment of adults with major depressive disorder: a systematic review and network meta-analysis. Lancet, 2018. 391(10128): p. 1357–1366.

11. BMJ, Large meta-analysis ends doubts about efficacy of antidepressants. 2018. k847.

12. Munkholm, K., A.S. Paludan-Muller, and K. Boesen, Considering the methodological limitations in the evidence base of antidepressants for depression: a reanalysis of a network meta-analysis. BMJ Open, 2019. 9(6): p. e024886.

13. Hamilton, M., Development of a rating scale for primary depressive illness. Br J Soc Clin Psychol, 1967. 6(4): p. 278–96.

14. Leucht, S., et al., What does the HAMD mean? J Affect Disord, 2013. 148(2-3): p. 243–8.

15. Guy, W., Clinical Global Impressions ECDEU Assessment Manual for Psychopharmacology, Revised (DHEW Publ. No. ADM 76-338). 1976, National Institute of Mental Health: Rockville, MD. p. 218–222.

16. Moncrieff, J. and I. Kirsch, Empirically derived criteria cast doubt on the clinical significance of antidepressant-placebo differences. Contemp Clin Trials, 2015. 43: p. 60–2.

17. BMJ, Effectiveness of antidepressants. 2018. k1073.

18. BMJ, The cost of dichotomising continuous variables. 2006. 1080.

19. Austin, P.C. and L.J. Brunner, Inflation of the type I error rate when a continuous confounding variable is categorized in logistic regression analyses. Stat Med, 2004. 23(7): p. 1159–78.

20. Senn, S., Statistical pitfalls of personalized medicine. Nature, 2018. 563(7733): p. 619–621.

21. Winkelbeiner, S., et al., Evaluation of Differences in Individual Treatment Response in Schizophrenia Spectrum Disorders: A Meta-analysis. JAMA Psychiatry, 2019.

22. Senn, S., Mastering variation: variance components and personalised medicine. Stat Med, 2016. 35(7): p. 966–77.

23. Fisher, R.A. and others, Statistical inference and analysis: Selected correspondence of ra fisher, edited by jh bennett. 1990: Oxford: Clarendon Press.

24. Montgomery, S.A. and M. Asberg, A new depression scale designed to be sensitive to change. Br J Psychiatry, 1979. 134: p. 382–9.

25. Rush, A.J., et al., Self-reported depressive symptom measures: sensitivity to detecting change in a randomized, controlled trial of chronically depressed, nonpsychotic outpatients. Neuropsychopharmacology, 2005. 30(2): p. 405–16.

26. Jefferson, J.W., et al., Extended-release bupropion for patients with major depressive disorder presenting with symptoms of reduced energy, pleasure, and interest: findings from a randomized, double-blind, placebo-controlled study. J Clin Psychiatry, 2006. 67(6): p. 865–73.

27. The Cochrane Collaboration, Cochrane Handbook for Systematic Reviews of Interventions Version 5.1.0. 2011 [02.09. 2019]; Available from: www.handbook.cochrane.org.

28. Nakagawa, S., et al., Meta-analysis of variation: ecological and evolutionary applications and beyond. Methods in Ecology and Evolution, 2015. 6(2): p. 143–152.

29. McCutcheon, R.A., et al., The efficacy and heterogeneity of antipsychotic response in schizophrenia: A meta-analysis. Mol Psychiatry, 2019. doi: 10.1038/s41380-019-0502-5.

30. Leucht, S., et al., Translating the HAM-D into the MADRS and vice versa with equipercentile linking. J Affect Disord, 2018. 226: p. 326–331.

31. Vehtari, A., A. Gelman, and J. Gabry. Practical Bayesian model evaluation using leave-one-out cross-validation and WAIC. 2016. doi: 10.1007/s11222-016-9696-4.

32. Volkmann, A. “A bound on the Treatment Effect Heterogeneity using estimates on the Variability Ratio”, in preparation.

33. Hengartner, M.P., Methodological Flaws, Conflicts of Interest, and Scientific Fallacies: Implications for the Evaluation of Antidepressants’ Efficacy and Harm. Front Psychiatry, 2017. 8: p. 275.

34. Jakobsen, J.C., C. Gluud, and I. Kirsch, Should antidepressants be used for major depressive disorder? BMJ Evid Based Med, 2019. doi: 10.1136/bmjebm-2019-111238.

35. Plöderl, M. and M.P. Hengartner, Can we expect that some patients respond better to antidepressants? A secondary variance-ratio meta-analysis. 2019. osf.io/98kex9.

36. Maslej, M.M., et al., Individual Differences in Response to Antidepressants: A Meta-analysis of Placebo-Controlled Randomized Clinical Trials. JAMA Psychiatry, 2020. DOI: 10.1001/jamapsychiatry.2019.4815

